# Estimation of SARS-CoV-2 aerosol emissions from simulated patients with COVID-19 and no to moderate symptoms

**DOI:** 10.1101/2020.04.27.20081398

**Authors:** Michael Riediker, Dai-Hua Tsai

## Abstract

**Importance:** Cases of the coronavirus disease 2019 (COVID-19) with no or mild symptoms were reported to frequently transmit the disease even without direct contact. The severe acute respiratory syndrome virus (SARS-COV-2) was found at very high concentrations in swab and sputum of such cases.

**Objective:** We aimed to estimate in a mathematical modeling study the virus release from such cases into different aerosol sizes by normal breathing and coughing, and what exposure can result from this in a room shared with such as case.

**Data Sources and Model:** We combined the size-distribution of exhaled breath microdroplets for coughing and normal breathing with viral sputum concentrations as approximation for lung lining liquid to obtain an estimate of emitted virus levels. The resulting emission data fed a single-compartment model of airborne concentrations in a room of 50 m^3^, the size of a small office or medical exam room.

**Results:** The estimated viral load in microdroplets emitted by simulated patients while breathing normally was on typical 0.0000049 copies/cm^3^ and could go up to 0.637 copies/cm^3^. The corresponding numbers for coughing simulated patients were 0.277 copies/cm^3^ and 36,030/cm^3^, respectively, per cough. The resulting concentrations in a room with a coughing emitter were always very high, up to 7.44 million copies/m^3^. However, also regular breathing microdroplets from high emitters was modelled to lead to 1248 copies/m^3^.

**Conclusions and Relevance:** In this modelling study, breathing and coughing were estimated to release large numbers of viruses, ranging from thousands to millions of virus copies/m^3^ in a room with an emitter having a high viral load, depending on ventilation and microdroplet formation process. These findings suggest that strict respiratory protection may be needed when there is a chance to be in the same room with a patient - whether symptomatic or not - especially for a prolonged time.

**Key Points:** *Question:* How much SARS-CoV-2 virus is released from a case by breathing and coughing, and what is the resulting concentration in a room?

*Finding:* In this mathematical modelling study, both, breathing and coughing were estimated to release large numbers of viruses, which can lead to millions of virus copies/m^3^ in a poorly ventilated room with a coughing emitter.

*Meaning:* These results may explain the important rate of transmissions and implies the need for strict respiratory protection when people are in the same room with a case with COVID-19.

## Introduction

The novel Coronavirus disease 2019 (COVID-19), emerged in late 2019 in Wuhan, China ^1^ from where it spread to the entire world. COVID-19 is caused by a novel type of Coronavirus, the severe acute respiratory syndrome virus (SARS-COV-2) ^2^. The host-receptor for SARS-CoV-2 was found to be Angiotensin I converting enzyme 2 (ACE2), which is present in cells of the lungs and airways ^3^. In the early phase of the outbreak, a large number of patients hospitalized for other reasons ^4^ and a considerable proportion of the medical staff ^5^ contracted COVID-19. However, the attack rate among medical staff corresponded to community rates when respiratory personal protective equipment (PPE) was used at work ^6,7^. Also a series of community-transmissions were reported from cases that had no apparent symptoms ^8–11^. The estimates for community and household attack rates are currently in the range of 1 % and 10 %, respectively ^12–15^. However, during super-spreading events in situations where many people engaged in loud voice activities gathered in closed rooms for prolonged time, such as a restaurant ^16^, a call-center ^17^, a dermatologists scientific board meeting ^18^, and a choir rehearsal ^19^ attack rates above 75% were reported. Notably, the choir rehearsal participants tried to follow social distancing and hand washing rules. These super-spreading events suggest that the airborne route may represent a virus transmission form in some indoor situations. Indeed, a study conducted in a Wuhan hospital found low airborne concentrations of the virus in the intensive care unit and in medical staff rooms ^20^. Correspondences about the viral load in samples from patients with COVID-19 having no or only mild symptoms reported very high concentrations of SARS-CoV-2 in samples taken in the nose, throat and saliva ^11,21–23^, and high during antiviral treatment ^24^. This all raised the question whether transfections could occur via the air.

When coughing, humans release thousands of microdroplets per cubic-centimeter in the size range of 0.6 to 15 µm, with the droplet concentration increasing strongly with cough flow rate ^25^. But also normal breathing will lead to some microdroplet production, which is attributed to fluid film rupture in the respiratory bronchioles during inhalation leading to the formation of droplets that are released during exhalation ^26^. The size of these droplets is mostly below 1 µm ^27^. The mode of droplet generation implies that they consist of lung lining liquid including dispersed viruses. Indeed, human volunteers exposed to virus-sized nanoparticles show nano-scaled particles in their exhaled breath ^28,29^. Also, the described size distribution of particles emitted from coughing as well as normal respiration suggests that an important proportion of them will be able to remain airborne for many hours in turbulent conditions ^30^.

## Objectives

This study aimed to estimate the cumulative viral load released from simulated patients with COVID-19 with no to moderate symptoms in different microdroplet sizes via respiration and coughing. We then used this information to make a risk appraisal for the situation of a low, typical or high emitter that is either breathing normally or coughing in a room operated at different air exchange rates. We chose a room size that is similar to a medical examination room or an office shared by two to three people.

## Design and Methods

### Concept

The release of viruses from individual simulated patients was modeled by first calculating the viral load per exhaled microdroplets formed during normal breathing and while coughing. The resulting size-distribution provided an initial estimate of the concentration of SARS-CoV-2 virus copies released by a regularly breathing or coughing simulated patient. This viral emission factor was then fed into a well-mixed one-compartment model to simulate the situation in a closed room with different ventilation air exchange rates. This study follows the concept of Strengthening The Reporting of Empirical Simulation Studies (STRESS) guideline ^31^. This mathematical modelling corresponds to a meta-analysis and was as such exempt from ethics approval.

### Data sources

Data on the number of viral copies present in sputum and swab samples were used to estimate the SARS-CoV-2 viral load present in the lining liquid of respiratory bronchioles in patients published before the here presented modelling (May 2020) ^11,21–24,32^, specifically 1,000 copies/ml representing a low-virus producing patient (“low emitter”), an “typical emitter” producing 10^6 copies/ml, and a “high emitter” producing 1*3^11 copies/ml. Exhaled microdroplet size distributions and numbers were retrieved from published studies on healthy persons coughing ^25^ and breathing normally ^26^. Both studies assessed the size-number distribution of freshly emitted microdroplets. The concentration of viral copies in each microdroplet size was calculated from the volume of the microdroplets, the actual count number in each size and the above-mentioned virus-load per ml sputum. The viral load in the actual microdroplet counts in each microdroplet size was then used to calculate the total viral concentration. The cumulative emissions in the PM10 fraction were summed up after applying the standard size fractionation curves ^33^ to the microdroplet distribution.

### Model

A one-compartment model ^34^ estimated the virus load concentration C for a perfectly mixed room of volume VR of 50 m^3^ with one simulated patient as source, using the following mass-balance (equation 1):

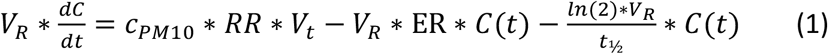

The emission rate was calculated from the concentration c_PM10_, the viral load in the PM_10_-size range, which are particles collected with a 50% efficiency cut-off at 10 µm aerodynamic diameter; and a respiratory rate of 15 breaths per minute (RR) at a tidal volume of V_t_ of 500 ml per breath. Air exchange rates (ER) used were 1-, 3-, 10-and 20-times per hour. The virus’ half-life t½ of 1.1 hours was obtained from an experimental study about the persistence of SARS-CoV-2 on surfaces and when airborne ^35^, tested by assessing the 50% tissue culture infective dose (TCID_50_).

The model for coughing was identical, except that coughing was assumed to happen every 30 seconds at a volume of 250 ml, as described for chronic dry cough patient (not having COVID-19) ^36^.

All statistics and models were calculated using Stata/SE 15.1 (Mac 64-bit Intel, Rev. 03 Feb 2020, StataCorp, College Station, TX, USA). Robust data reported include estimated averages and ranges. The models and code are available on request.

## Results

### Emissions from normal breathing simulated patients

To estimate the virus emissions from simulated patients breathing normally, we first calculated the viral load for the microdroplet size distribution. Figure 1 shows that the highest virus load is present in the largest microdroplet size. The cumulative total emission per breath was 0.0000000049 copies/cm^3^(air) for a low emitter, 0.0000049 copies/cm^3^ for an typical simulated patient, and 0.637 copies/cm^3^ for a high emitter. The cumulative emissions in the PM_10_ fraction were approximately 1/3 of these values with 0.0000017 copies/cm^3^ (typical) and 0.226 copies/cm^3^ (high) per breath.

**Figure 1:**
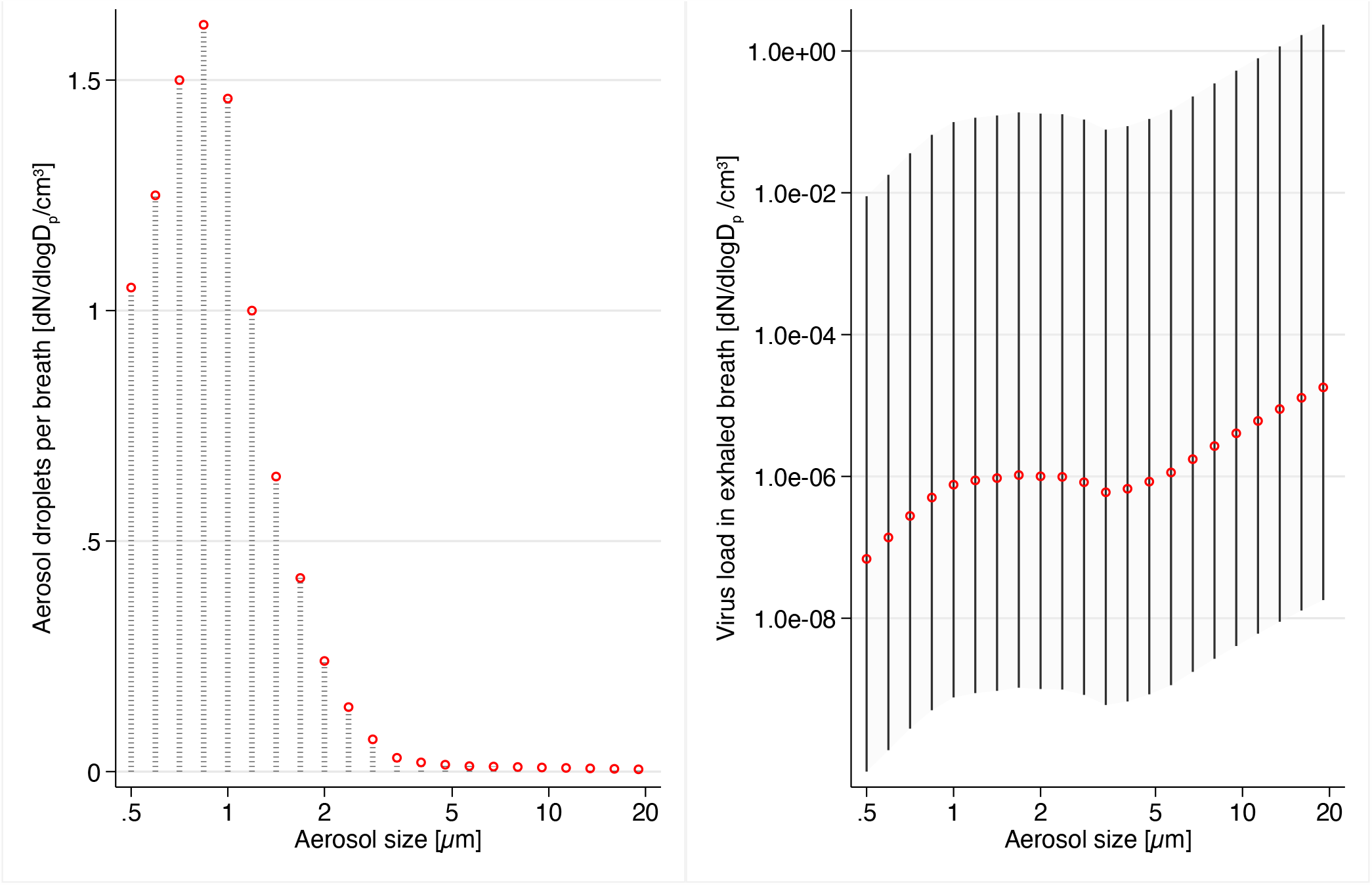
Size distribution of exhaled microdroplets (left) and resulting viral emissions (right) during normal breathing. The left panel shows the typical exhaled microdroplet concentration used as input for the simulation, the right panel shows the modelled viral emission per breath for typical (red), high and low emitters (spike-lines).

### Emission from coughing simulated patient

We then estimated the virus emissions from a coughing simulated patient (Figure 2). The cumulative total emission per cough was 0.000277 copies/cm^3^ for a low emitter, 0.277 copies/cm^3^ for an typical simulated patient, and 36,030 copies/cm^3^ for a high emitter. The cumulative emissions in the PM_10_ fraction were about 1/2 of these values with 0.156 copies/cm^3^ (typical) and 20,221 copies/cm^3^ (high) per cough.

**Figure 2:**
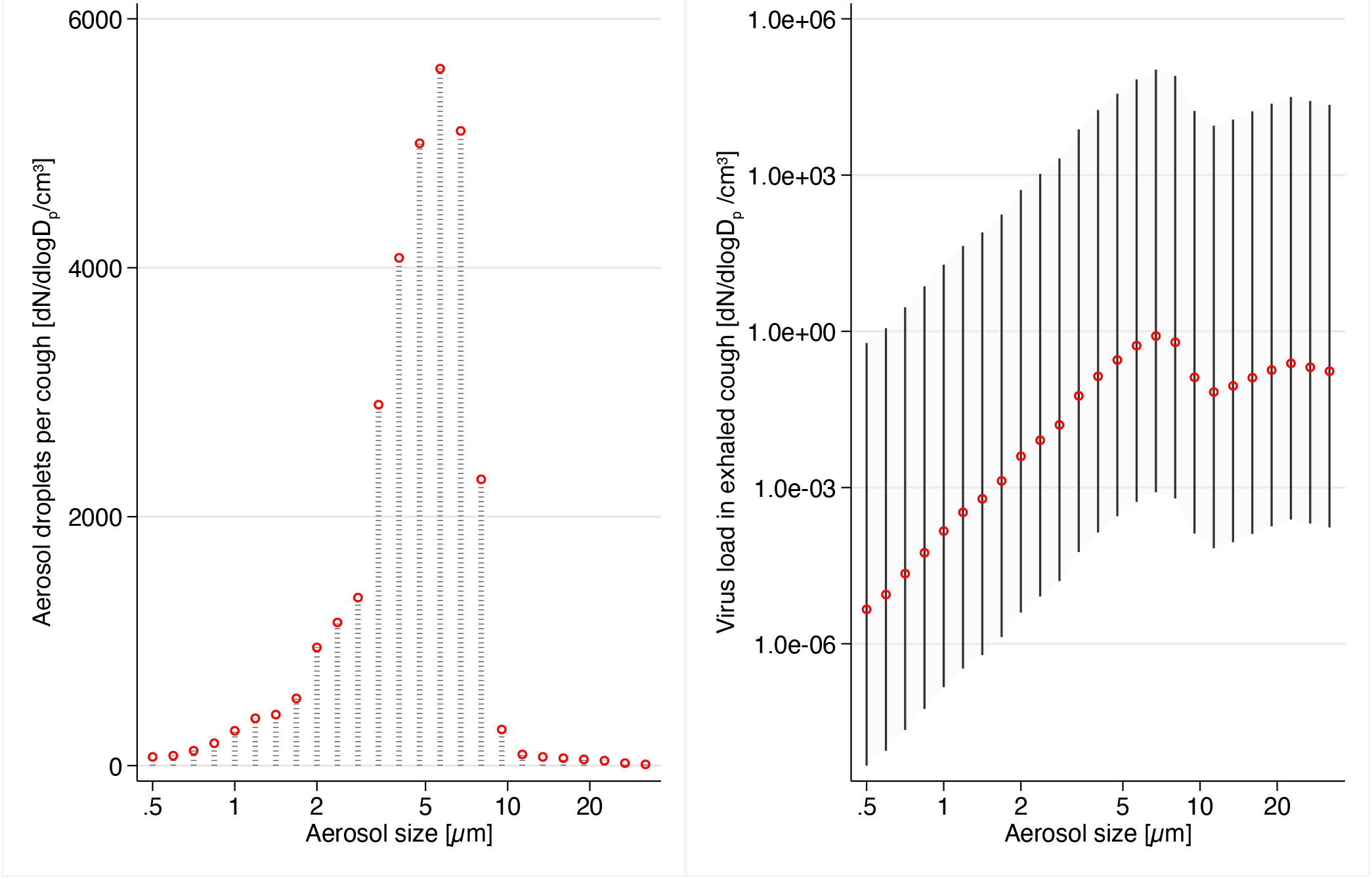
Size distribution of exhaled microdroplets (left) and resulting viral emissions (right) during coughing. The left panel shows the typical exhaled microdroplet concentration used as input for the simulation, the right panel shows the modelled viral emission per breath for typical (red), high and low emitters (spike-lines).

### Exposure estimation for bystanders

To estimate the exposure of bystanders spending time in the same room as a person with COVID-19, we calculated the time-course of the viral load in the thoracic size fraction for small droplets released from a high-emitter either breathing normally or coughing. Figure 3 shows the results for a high-emitting simulated patient coughing frequently.

**Figure 3:**
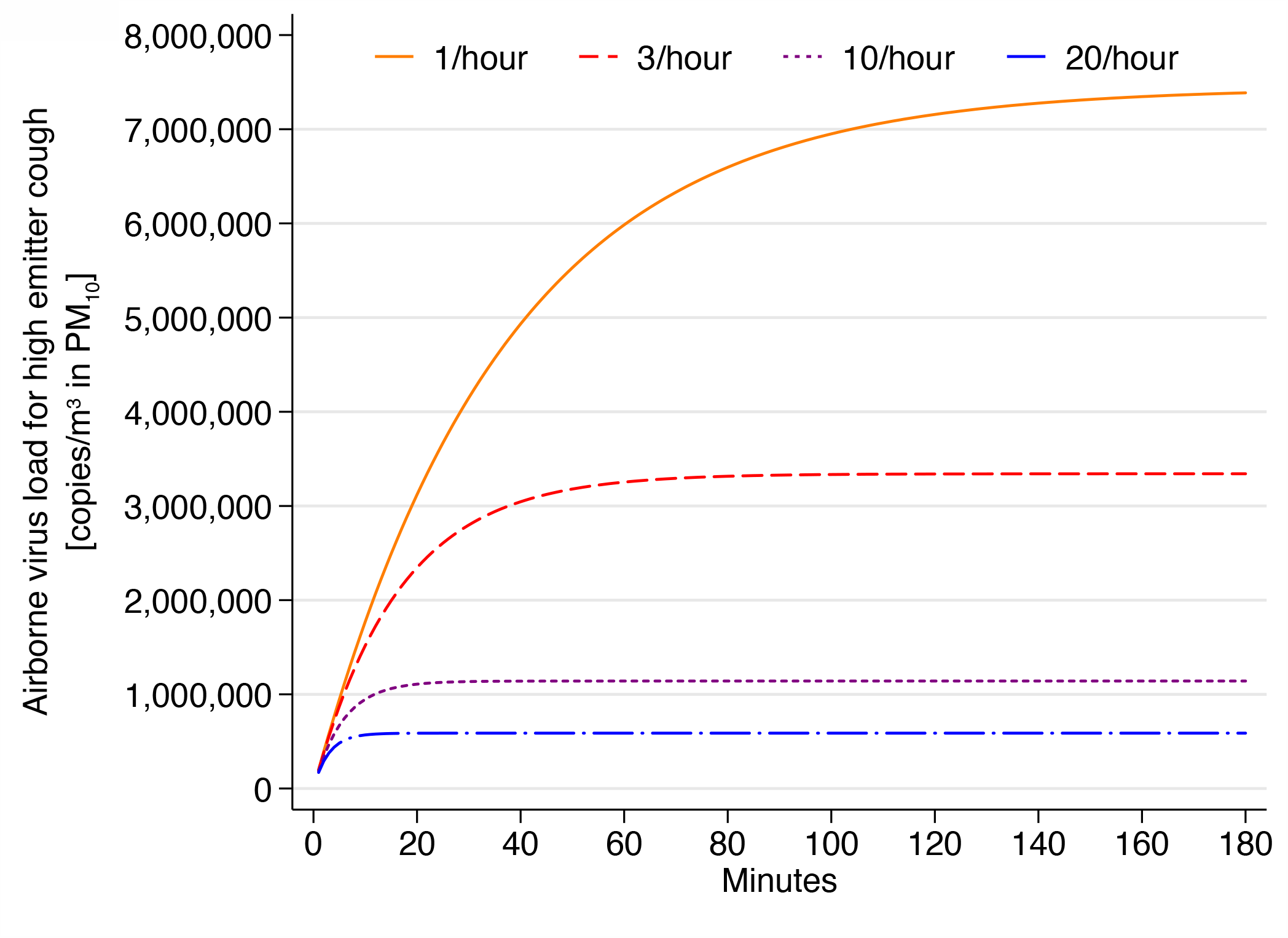
Temporal course of airborne virus load in a perfectly mixed room of 50 m^3^. The simulation estimated the concentration in a closed room for different air exchange rates. The emitter was assumed to have a high virus-load in the lungs and to be coughing intermittently every 30 seconds.

For a typical hospital ventilation situation of 10 air exchanges per hour, the concentration plateaus after about 30 minutes, while for a typical office with 3 air exchanges/hour, concentrations continue to rise for over one hours. In the used model, concentrations scale linearly with the simulated patient emission rate, the plateau concentrations for different emitting simulated patients and ventilation types are summarized in Table 1.

**Table 1:** Plateau concentration for different combinations of air exchange rate, emission form and emitter type.

| Air exchange rate (times / hour) | 1 / hour | 3 / hour | 10 / hour | 20 / hour |
| --- | --- | --- | --- | --- |
| Time until 99% of plateau | 169 minutes | 77 minutes | 26 minutes | 14 minutes |
| Airborne viral concentration at plateau (copies/m<sup>3</sup>) |  |  |  |  |
| Regular breathing |  |  |  |  |
| Low emitter | 0.00000960 | 0.00000431 | 0.00000147 | 0.00000076 |
| Typical emitter | 0.00960 | 0.00431 | 0.00147 | 0.00076 |
| High emitter | 1247.7 | 560.3 | 191.3 | 98.6 |
| Frequent coughing (every 30 seconds) |  |  |  |  |
| Low emitter | 0.5725 | 0.2571 | 0.00878 | 0.00452 |
| Typical emitter | 57.250 | 25.709 | 8.779 | 4.524 |
| High emitter | 7 442 598 | 3 342 148 | 1 141 326 | 588 093 |

## Discussion

An elevated number of viruses is expected to be released by patients with COIVD-19 having high viral load in the form of airborne microdroplets, especially when they are coughing. While the bigger portion of the emitted viral load is in the form of large droplets that can deposit rapidly, there is also an important portion in the smaller size fractions. Small microdroplets can remain airborne for an extended time ^30^ and are very effective at reaching the lungs ^37^.

One study assessed airborne SARS-CoV-2 levels in a hospital in Wuhan, China and found concentrations in the range of 20 copies/m^3^ in medical staff offices and meeting rooms ^20^, concentrations that our modelling would suggest for a small room with a regularly breathing non-symptomatic person having a viral load slightly above a typical emitter.

An typical person breathes about a half m^3^ per hour in resting state ^38^, which can rapidly increase to several m^3^ during exercise ^39^. Thus, a person spending time in a room with an typical emitting patient breathing normally has the chance of inhaling only a few copies of the virus when keeping distance from that person. However, the situation is worse in the presence of a high emitter and worst if the patient is a coughing high emitter. A review of a wide range of respiratory viruses suggests that the infective dose is often quite low. Sometimes as few as few hundred units of active virus (TCID_50_) ^40^ seem sufficient to provoke a disease. Thus, our modelling suggests that there is a clear risk of infections for a person spending an extended time in the room with an infected person having an elevated viral load, even if the distance is too large for direct transmission. The situation is worse if the person is coughing.

High emitters are not very frequent in the population. However, if such a person is engaged in activities such as loud speaking or singing, microdroplet formation and thus viral emissions can rapidly increase by one to two orders of magnitude ^41^. This may help explain the occasional superspreading events in crowded situations involving loud voices ^16–19^.

The occasionally very high virus load in exhaled respiratory microdroplets proposed by our assessment may be an explanation why COVID-19 was associated with more transfections to hospital staff than what was expected from SARS ^4^. While having everybody wear a surgical face mask can be an effective source control ^42^, the protective factors may still be insufficient if an extended amount of time is spent in the same room with a coughing high emitter, especially if the room is small and the ventilation low. Increasing ventilation can help to some extent but is not sufficient in a room of the size of a typical office or medical exam room. Note also that ventilation design for hospitals is complex and not always functioning as intended ^43^.

The implications for the normal life and the workplace are that the risk of infection is real when being near an infected person with high viral load in a room for more than a few minutes and this even when keeping distance to that person. Sharing a workplace in a small room with a non-symptomatic case seems not advised. This implies that workplaces should not be shared as long as there are no rapid tests to differentiate between healthy and non-symptomatic cases. Medical staff is advised to wear the best possible respiratory protection whenever in the same room as a patient, especially when this person is coughing, in which case eye protection is advised as well ^44^. In addition, every patient, also non-symptomatic ones, should wear a well-fitting surgical face mask to reduce emissions, which will increase the overall protection for the medical staff ^42^.

## Limitations

Our assessment has a number of limitations. Namely: 1) The estimated virus levels strongly depend on the number of virus copies produced by a case with COVID-19. We used sputum data from a well described peer-reviewed study ^21^ assuming that it is a reasonable approximation for the virus load in the respiratory bronchioles, the space where most respiratory microdroplets are formed. Our high-emitter estimates would be 100-fold higher if the most extreme viral data was combined with microdroplet super-emissions ^22,41^. 2) We used information about virus copies but compare the results with TCID_50_ infective dose. Research on other virus types suggests that the number of virus copies and TCID_50_ are comparable ^45^. However, it would be important to confirm this relationship for the case of SARS-CoV-2. 3) For breath and cough microdroplets release, we used data collected in experimental setups involving healthy young subjects. However, microdroplet formation is influenced by surface tension of the lung lining liquid ^46^. It is likely that microdroplet formation will be altered in cases with COVID-19 but it is not clear in which direction. 4) Microdroplets will shrink in dry air ^47^, resulting in a shift to smaller particle sizes. This will not directly change the number of copies in the PM_10_ range but simply upconcentrate the viral load per microdroplet. While we addressed passivation of viruses in the air by using the documented half-life ^35^, it is still possible that viruses in smaller droplets are quicker passivated because of shorter diffusion distances for airborne oxidants and faster increasing salinity. Our estimates would be slightly smaller if this was relevant. 5) The one-compartment model assumes perfectly mixed conditions. However, often, rooms are not perfectly mixed and also ventilation and room geometry will add spatiotemporal variability. The modelling provides an estimate, but exact concentrations will vary in function of the real circumstances. In multi-room situations, numerical flow simulations seem indicated to describe the microdroplet distribution ^48^. 6) Finally, though our results suggest that in certain situations, airborne transmission of COVID-19 may be possible, it is important to keep in mind that this was a modelling effort. While this route would provide a convenient explanation for several superspreading events ^16–19^, and even though the virus was found in airborne microdroplets in hospital situations ^20^, it still needs to be validated in clinical settings and animal models.

## Conclusions

In conclusion, our mathematical modelling suggests that the viral load in the air can rapidly reach critical concentrations in small and ill-ventilated rooms, especially when the patient is a super-spreader defined as a person emitting large number of microdroplets containing a high viral load. Thus, strict respiratory protection is needed whenever there is a chance to be in the same room with such a patient - whether symptomatic or not - especially if this was for a prolonged time.

## Data Availability

All data sources are mentioned in the manuscript and publicly available.

## Acknowledgments

This modelling was entirely funded by the Swiss Centre for Occupational and Environmental Health (SCOEH). No external funder had any role in the design and conduct of the study; collection, management, analysis, and interpretation of the data; preparation, review, or approval of the manuscript; and decision to submit the manuscript for publication. Both authors had full access to all the data in the study and take responsibility for the integrity of the data and the accuracy of the data analysis.

## References

1. Zhu N, Zhang D, Wang W, et al. A Novel Coronavirus from Patients with Pneumonia in China, 2019. N Engl J Med. 2020;382(8):727–733. doi:10.1056/NEJMoa2001017

2. Gorbalenya AE, Baker SC, Baric RS, et al. Severe Acute Respiratory Syndrome-Related Coronavirus: The Species and Its Viruses – a Statement of the Coronavirus Study Group. Microbiology; 2020. doi:10.1101/2020.02.07.937862

3. Qi F, Qian S, Zhang S, Zhang Z. Single cell RNA sequencing of 13 human tissues identify cell types and receptors of human coronaviruses. Biochem Biophys Res Commun. Published online March 2020:S0006291×20305234. doi:10.1016/j.bbrc.2020.03.044

4. Wang D, Hu B, Hu C, et al. Clinical Characteristics of 138 Hospitalized Patients With 2019 Novel Coronavirus–Infected Pneumonia in Wuhan, China. JAMA. 2020;323(11):1061. doi:10.1001/jama.2020.1585

5. Chu J, Yang N, Wei Y, et al. Clinical characteristics of 54 medical staff with COVID-19: A retrospective study in a single center in Wuhan, China. J Med Virol. Published online April 6, 2020:jmv.25793. doi:10.1002/jmv.25793

6. Kluytmans-van den Bergh MFQ, Buiting AGM, Pas SD, et al. Prevalence and Clinical Presentation of Health Care Workers With Symptoms of Coronavirus Disease 2019 in 2 Dutch Hospitals During an Early Phase of the Pandemic. JAMA Netw Open. 2020;3(5):e209673. doi:10.1001/jamanetworkopen.2020.9673

7. Cheng VC-C, Wong S-C, Yuen K-Y. Estimating Coronavirus Disease 2019 Infection Risk in Health Care Workers. JAMA Netw Open. 2020;3(5):e209687. doi:10.1001/jamanetworkopen.2020.9687

8. Bai Y, Yao L, Wei T, et al. Presumed Asymptomatic Carrier Transmission of COVID-19. JAMA. Published online February 21, 2020. doi:10.1001/jama.2020.2565

9. Kimball A, Hatfield KM, Arons M, et al. Asymptomatic and Presymptomatic SARS-CoV-2 Infections in Residents of a Long-Term Care Skilled Nursing Facility - King County, Washington, March 2020. MMWR Morb Mortal Wkly Rep. 2020;69(13):377–381. doi:10.15585/mmwr.mm6913e1

10. Li R, Pei S, Chen B, et al. Substantial undocumented infection facilitates the rapid dissemination of novel coronavirus (SARS-CoV2). Science. Published online March 16, 2020. doi:10.1126/science.abb3221

11. Zou L, Ruan F, Huang M, et al. SARS-CoV-2 Viral Load in Upper Respiratory Specimens of Infected Patients. N Engl J Med. 2020;382(12):1177–1179. doi:10.1056/NEJMc2001737

12. Cheng H-Y, Jian S-W, Liu D-P, et al. Contact Tracing Assessment of COVID-19 Transmission Dynamics in Taiwan and Risk at Different Exposure Periods Before and After Symptom Onset. JAMA Intern Med. Published online May 1, 2020. doi:10.1001/jamainternmed.2020.2020

13. Bi Q, Wu Y, Mei S, et al. Epidemiology and transmission of COVID-19 in 391 cases and 1286 of their close contacts in Shenzhen, China: a retrospective cohort study. Lancet Infect Dis. Published online April 2020:S1473309920302875. doi:10.1016/S1473-3099(20)30287-5

14. Li W, Zhang B, Lu J, et al. The characteristics of household transmission of COVID-19. Clin Infect Dis. Published online April 17, 2020 :ciaa450. doi:10.1093/cid/ciaa450

15. COVID-19 National Emergency Response Center, Epidemiology and Case Management Team, Korea Centers for Disease Control and Prevention. Coronavirus Disease-19: Summary of 2,370 Contact Investigations of the First 30 Cases in the Republic of Korea. Osong Public Health Res Perspect. 2020;11(2):81–84. doi:10.24171/j.phrp.2020.11.2.04

16. Lu J, Gu J, Li K, et al. COVID-19 Outbreak Associated with Air Conditioning in Restaurant, Guangzhou, China, 2020. Emerg Infect Dis. 2020;26(7). doi:10.3201/eid2607.200764

17. Shin Young Park, Young-Man Kim, Seonju Yi, et al. Coronavirus Disease Outbreak in Call Center, South Korea. Emerg Infect Dis J. 2020;26(8). doi:10.3201/eid2608.201274

18. Hijnen D, Marzano AV, Eyerich K, et al. SARS-CoV-2 Transmission from Presymptomatic Meeting Attendee, Germany. Emerg Infect Dis. 2020;26(8). doi:10.3201/eid2608.201235

19. Hamner L, Dubbel P, Capron I, et al. High SARS-CoV-2 Attack Rate Following Exposure at a Choir Practice — Skagit County, Washington, March 2020. MMWR Morb Mortal Wkly Rep. 2020;69(19):606–610. doi:10.15585/mmwr.mm6919e6

20. Liu Y, Ning Z, Chen Y, et al. Aerodynamic analysis of SARS-CoV-2 in two Wuhan hospitals. Nature. Published online April 27, 2020. doi:10.1038/s41586-020-2271-3

21. Wölfel R, Corman VM, Guggemos W, et al. Virological assessment of hospitalized patients with COVID-2019. Nature. Published online April 1, 2020. doi:10.1038/s41586-020-2196-x

22. Pan Y, Zhang D, Yang P, Poon LLM, Wang Q. Viral load of SARS-CoV-2 in clinical samples. Lancet Infect Dis. 2020;20(4):411–412. doi:10.1016/S1473-3099(20)30113-4

23. Pasomsub E, Watcharananan SP, Boonyawat K, et al. Saliva sample as a non-invasive specimen for the diagnosis of coronavirus disease-2019 (COVID-19): a cross-sectional study. Clin Microbiol Infect. Published online May 2020:S1198743×20302780. doi:10.1016/j.cmi.2020.05.001

24. Zheng S, Fan J, Yu F, et al. Viral load dynamics and disease severity in patients infected with SARS-CoV-2 in Zhejiang province, China, January-March 2020: retrospective cohort study. BMJ. Published online April 21, 2020:m1443. doi:10.1136/bmj.m1443

25. Yang S, Lee GWM, Chen C-M, Wu C-C, Yu K-P. The Size and Concentration of Droplets Generated by Coughing in Human Subjects. J Aerosol Med. 2007;20(4):484–494. doi:10.1089/jam.2007.0610

26. Johnson GR, Morawska L. The Mechanism of Breath Aerosol Formation. J Aerosol Med Pulm Drug Deliv. 2009;22(3):229–237. doi:10.1089/jamp.2008.0720

27. Papineni RS, Rosenthal FS. The Size Distribution of Droplets in the Exhaled Breath of Healthy Human Subjects. J Aerosol Med. 1997;10(2):105–116. doi:10.1089/jam.1997.10.105

28. Gschwind S, Graczyk H, Günther D, Riediker M. A method for the preservation and determination of welding fume nanoparticles in exhaled breath condensate. Env Sci Nano. 2016;3(2):357–364. doi:10.1039/C5EN00240K

29. Sauvain J-J, Hohl MSS, Wild P, Pralong JA, Riediker M. Exhaled breath condensate as a matrix for combustion-based nanoparticle exposure and health effect evaluation. J Aerosol Med Pulm Drug Deliv. 2014;27(6):449–458. doi:10.1089/jamp.2013.1101

30. Willeke K, Baron PA, eds. Aerosol Measurement: Principles, Techniques, and Applications. Van Nostrand Reinhold; 1993.

31. Monks T, Currie CSM, Onggo BS, Robinson S, Kunc M, Taylor SJE. Strengthening the reporting of empirical simulation studies: Introducing the STRESS guidelines. J Simul. 2019;13(1):55–67. doi:10.1080/17477778.2018.1442155

32. The COVID-19 Investigation Team. Clinical and virologic characteristics of the first 12 patients with coronavirus disease 2019 (COVID-19) in the United States. Nat Med. Published online April 23, 2020. doi:10.1038/s41591-020-0877-5

33. BSI. EN 481:1993 Workplace Atmospheres. Size Fraction Definitions for Measurement of Airborne Particles. BSI; 1993.

34. Scheff PA, Paulius VK, Curtis L, Conroy LM. Indoor Air Quality in a Middle School, Part II: Development of Emission Factors for Particulate Matter and Bioaerosols. Appl Occup Environ Hyg. 2000;15(11):835–842. doi:10.1080/10473220050175715

35. van Doremalen N, Bushmaker T, Morris DH, et al. Aerosol and Surface Stability of SARS-CoV-2 as Compared with SARS-CoV-1. N Engl J Med. Published online March 17, 2020:NEJMc2004973. doi:10.1056/NEJMc2004973

36. Hsu JY, Stone RA, Logan-Sinclair RB, Worsdell M, Busst CM, Chung KF. Coughing frequency in patients with persistent cough: assessment using a 24 hour ambulatory recorder. Eur Respir J. 1994;7(7):1246–1253. doi:10.1183/09031936.94.07071246

37. Lippmann M, Yeates DB, Albert RE. Deposition, retention, and clearance of inhaled particles. Br J Ind Med. 1980;37(4):337–62.

38. Beardsell I, ed. Get through MCEM. Part A: MCQs. RSM; 2011.

39. Ramos CA, Reis JF, Almeida T, Alves F, Wolterbeek HT, Almeida SM. Estimating the inhaled dose of pollutants during indoor physical activity. Sci Total Environ. 2015;527-528:111–118. doi:10.1016/j.scitotenv.2015.04.120

40. Yezli S, Otter JA. Minimum Infective Dose of the Major Human Respiratory and Enteric Viruses Transmitted Through Food and the Environment. Food Environ Virol. 2011;3(1):1–30. doi:10.1007/s12560-011-9056-7

41. Asadi S, Wexler AS, Cappa CD, Barreda S, Bouvier NM, Ristenpart WD. Aerosol emission and superemission during human speech increase with voice loudness. Sci Rep. 2019;9(1):2348. doi:10.1038/s41598-019-38808-z

42. Patel RB, Skaria SD, Mansour MM, Smaldone GC. Respiratory source control using a surgical mask: An in vitro study. J Occup Environ Hyg. 2016;13(7):569–576. doi:10.1080/15459624.2015.1043050

43. Grosskopf KR, Herstein KR. The aerodynamic behavior of respiratory aerosols within a general patient room. 2012;18(4):15.

44. Chen M-J, Chang K-J, Hsu C-C, Lin P-Y, Liu CJ-L. Precaution and Prevention of Coronavirus Disease 2019 (COVID-19) Infection in the Eye: J Chin Med Assoc. Published online April 2020:1. doi:10.1097/JCMA.0000000000000334

45. Kim J-O, Kim W-S, Kim S-W, et al. Development and Application of Quantitative Detection Method for Viral Hemorrhagic Septicemia Virus (VHSV) Genogroup IVa. Viruses. 2014;6(5):2204–2213. doi:10.3390/v6052204

46. Edwards DA, Man JC, Brand P, et al. Inhaling to mitigate exhaled bioaerosols. Proc Natl Acad Sci. 2004;101(50):17383–17388. doi:10.1073/pnas.0408159101

47. Holmgren H, Bake B, Olin A-C, Ljungström E. Relation Between Humidity and Size of Exhaled Particles. J Aerosol Med Pulm Drug Deliv. 2011;24(5):253–260. doi:10.1089/jamp.2011.0880

48. Chang T-J, Hsieh Y-F, Kao H-M. Numerical investigation of airflow pattern and particulate matter transport in naturally ventilated multi-room buildings. Indoor Air. 2006;16(2):136–152. doi:10.1111/j.1600-0668.2005.00410.x

